# SARS-CoV-2 Antibody Formation Among Healthcare Workers, September 2, 2020

**DOI:** 10.1101/2020.09.10.20192104

**Authors:** Thomas Birch, Ravit Barkama, Joanna Tyszkiewicz Georgescu, Emma Yamada, Drew Olsen, Ed Torres, Alison Sinclair

## Abstract

Many frontline healthcare workers throughout the world have been exposed to COVID-19 infection in the workplace and the community. We describe the nature of infection and the durability of antibodies among various types of healthcare workers at an acute care community hospital in northern New Jersey adjacent to New York City, part of the epicenter of the first wave of the US epidemic. Exposure was concentrated among frontline workers and in clusters among support staff. The antibody response correlated with symptoms and job type.

**Methods:** Employees had Polymerase Chain Reaction testing using a variety of commercially available products based on availability. All screening and testing for diagnosis used the nasopharyngeal swab. Tests for SARS-CoV-2 were obtained at approximately 2, 3 and 4 months using Abbott IgG antibody assay. Results of community sero-prevalence were obtained from hospital and physician surveys as no government entity was monitoring sero-prevalence. Hospital associated employee COVID-19 infections were identified through testing and contact tracing.

**Results:** 5179 patients with COVID-19 syndrome were managed through telehealth over 6 months. 3236 patients were admitted with COVID-19 disease during this time. 2514 out of 3100 employees were tested for antibody to SARS-CoV-2. Overall, 16% of employees tested positive for antibodies at some point. When divided by general job type, 17% of direct caregivers were positive. 16% of indirect contact such as receptionists, housekeeping and food service workers were positive. 16% of nonclinical workers with no patient contact tested positive. The community rate of sero-positivity in one study in Bergen County was 12.2. (1) In the midst of the epidemic, the rate among hospital workers in New York City was 13.7%. (2) The overall population rate in New York City, at the peak of the epidemic, was 21% with some communities as high as 68%. Long Island had 16.7% and Westchester/Rockland 11.7%. (3) Other hospital systems in Northern NJ had up to 25% sero-positive employees. (4)

---

The rate of positivity for departments, with more than 10 employees resulted, ranges from 3.5% in Diagnostic Radiology to 52% in Psychiatry. Notable high rates include: ER Registration 44%, ER Staff 24%, Med-Surg Nursing Units 29 to 38% and Security 31%. Other frontline staff such as Physical Therapy 30% and Respiratory Therapy 33% also had high rates of exposure. Notable low rates included: Labor and Delivery 11%, Preadmission Testing 8.3%, Pharmacy 6.4% and Operating Room staff 4.4%.

A sample of the first 200 employees with COVID-19 symptoms were divided into six groups.

| <u>COVID-19 Syndrome</u> | <u>Nasopharyngeal PCR</u> | <u>SARS-CoV-2 Antibody</u> |
| --- | --- | --- |
| Positive | Positive | 96% |
| Positive | Negative, overall | 18% |
| Positive | Negative had fever | 48% |
| Positive | Negative without fever | 7% |
| Positive | Not tested | 17% |
| Negative (control) | Not tested | 3% |

Time course of antibody responses for 56 participants with or without fever.

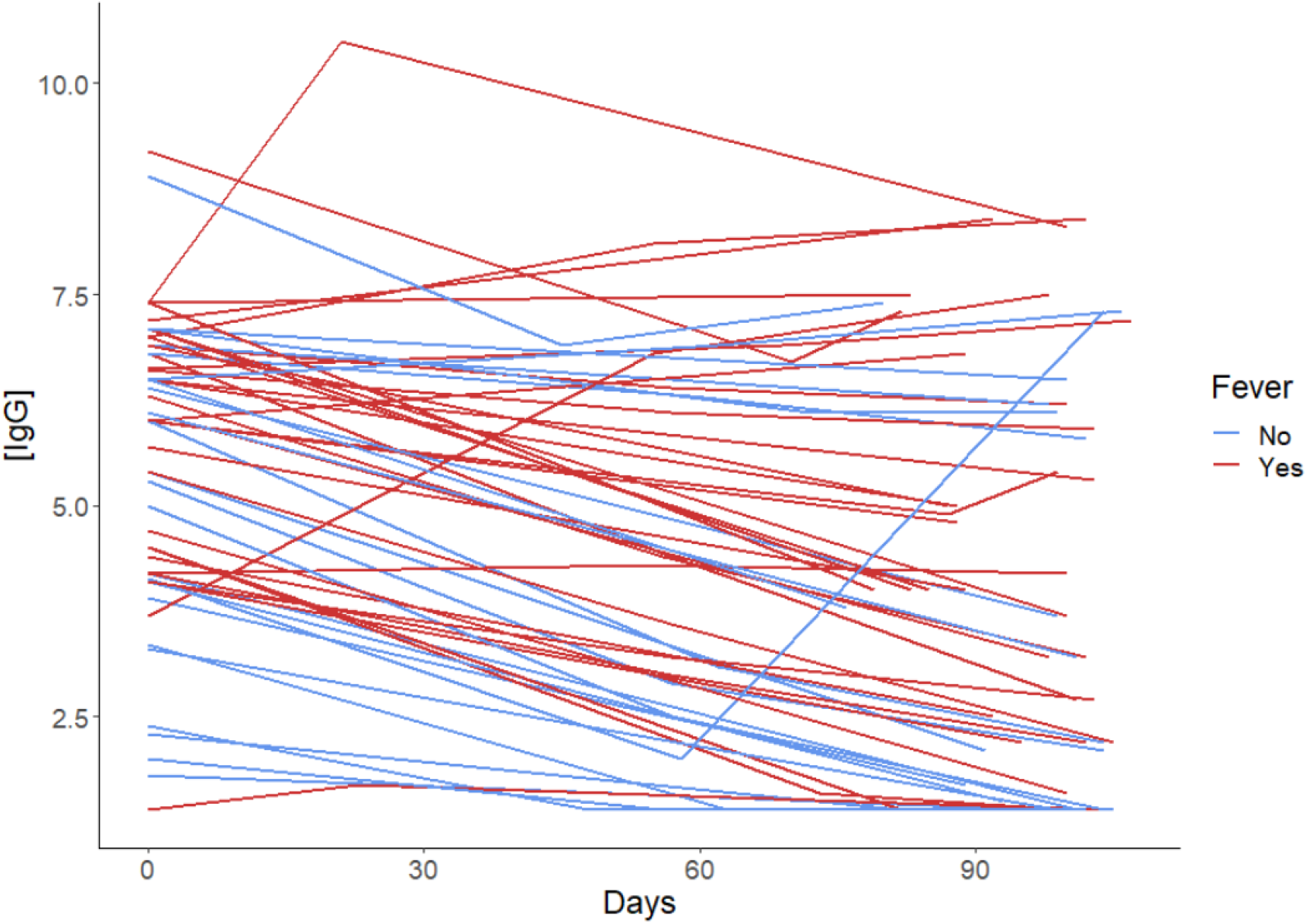

Time course of normalized antibody levels for 56 participants with or without fever.

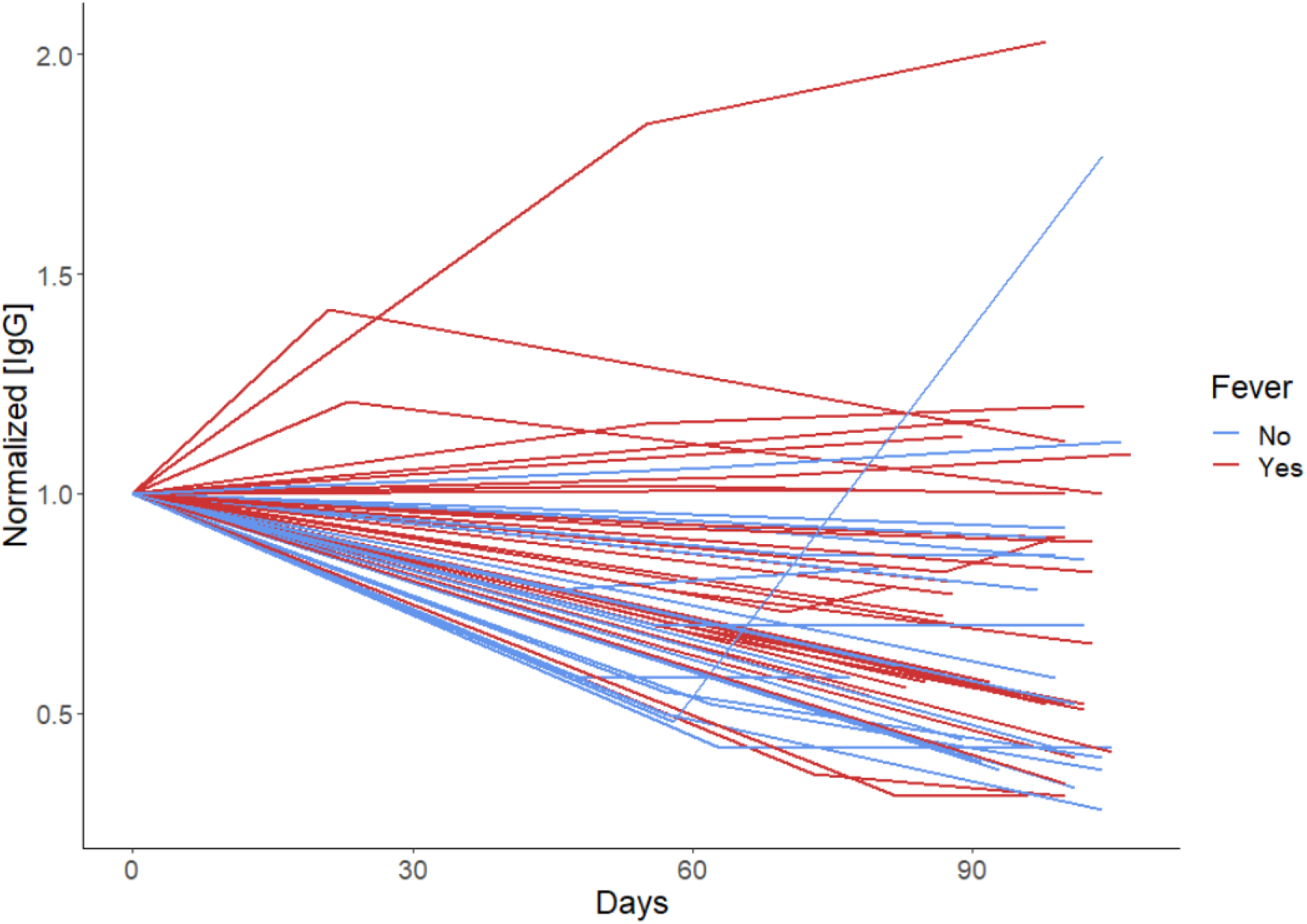

Log base normalization of [IgG] was used to linearize the predictor-target relationship and antibody half-life was calculated using correlation coefficients. The antibody half-life of 148 days.

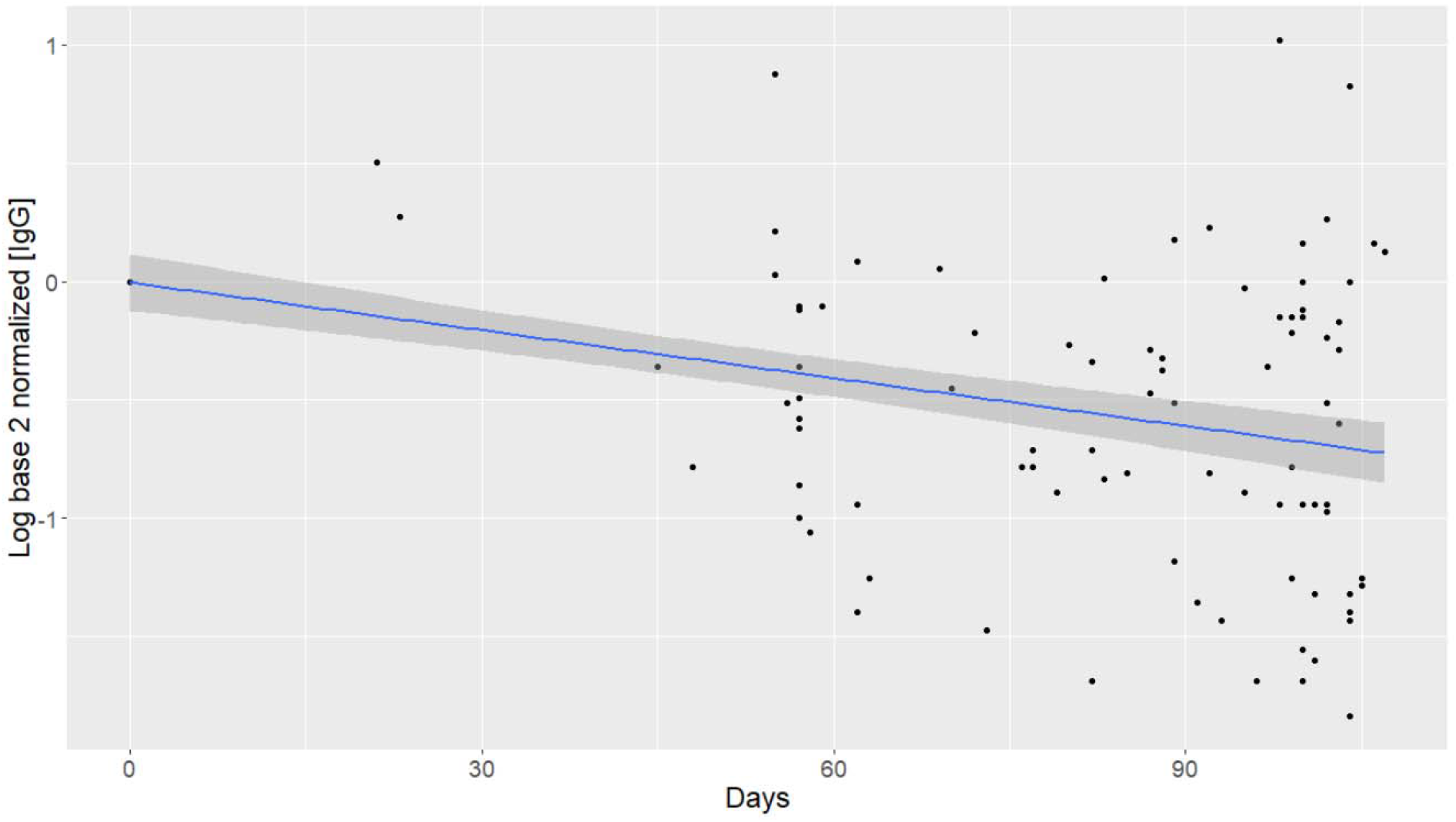

## Interpretation

For most employees, it is not possible to determine whether they acquired infection in the hospital or in the community. When the rate for job classifications is much higher than the background community rate or when contact tracing yields a likely exposure for a group of individuals, the site of transmission can often be inferred. Early on, there was an excessive focus on the travel history from Asia and specifically China to identify risk. It was unrecognized that the pandemic had entered the New York City area from Europe and local transmission in our communities was rapidly accelerating. As a result, many staff were exposed without Personal Protective Equipment at that time. It was not uniform policy for all employees to wear masks when gathering outside of patient care areas such as the cafeteria, break rooms and conference rooms. There was no hospital associated infections transmitted to employees after April 15^th^ when the true character of the epidemic was realized and policies were improved.

Many employees had a COVID-19 syndrome with exposure in the midst of the epidemic but had negative nasopharyngeal swabs. This is likely due to the absence of the virus in the nasopharynx even as they had infection and went on to make antibodies. It is unlikely to be operator error in swabbing because a small cadre of trained technicians and nurses performed all of the swabbing. Many more had a mild COVID-19 syndrome with exposure in the midst of the epidemic and likely had true SARS-CoV-2 infection but did not make the antibody measured in this assay especially if they did not have fever. Fever appears to be a very important indicator of antibody response because it is an indicator of more severe and systemic disease. Milder and more local disease may be due to exposure to a low level inoculum. The severity of illness in COVID-19 has been shown to correlate with the use of face masks which likely reduce inoculum. (5) There were clusters of COVID-19 cases in Administration, Information Systems and Plant Maintenance which likely originated from the community with subsequent intrahospital spread.

There was an abrupt drop in hospital acquired COVID-19 infections among employees when the true sources and modes of infection were discovered. Among physicians, primary care doctors with unprotected exposures in their offices were disproportionately affected while all 9 critical care specialists with extensive but protected exposure in the ICU, remained uninfected. This demonstrates that Personal Protective Equipment is effective in preventing infection and it is the unrecognized infected and often asymptomatic or minimally symptomatic coworker or patient who represents the greatest risk for transmission.

The overall half-life for the SARS-CoV-2 IgG antibody measured with multiple determinations was 148 days with confidence range 131 to 171 days which is consistent with serologic responses after many viral infections. It is significantly longer than the 36 days, measured on two occasions, in a cohort of community patients. (6) Many had stable or rising antibody levels for unclear reasons.

Most theorists estimate that 16% immunity is inadequate to prevent a resurgence of infection in an uncontrolled environment. It is not possible to factor in the effect of careful mask wearing, hand hygiene and disinfection but these activities coupled with an additional increment of unmeasured immunity among those with infections too mild to elicit antibody will likely be sufficient to protect healthcare workers through any community resurgence that may occur in the fall or winter. Further research is needed into the full spectrum and duration of immunity to SARS-CoV-2.

## Data Availability

(1) bergencountyhealth.gov Bergen County Mobile Testing Statistics and Locations July 2, 2020
(2) Moscola J et al. Prevalence of SARS-CoV-2 Antibodies in Health Care Personnel in New York City Area. JAMA. 2020; 324:893-5. DOI:10.1001
(3) Goldstein J. 1.5 Million Tests Show What Parts of NYC Were Hit Hardest. NYT, Aug 19,2020
(4) Personal communication
(5) Gandi M et al. Masks do more than protect others during COVID-19: reducing the inoculum of SARS-CoV-2 to protect the wearer. J Gen Intern Med 2020July 31 (Epub ahead of print)
(6) Ibarrondo JF et al. Rapid Decay of Anti-SARS-CoV-2 Antibodies in Persons with Mild Covid-19. NEJM 2020 Jul 21, DOI: 10.1056

